# Effectiveness of airport screening at detecting travellers infected with 2019-nCoV

**DOI:** 10.1101/2020.01.31.20019265

**Authors:** Billy Quilty, Sam Clifford, CMMID nCoV working group, Stefan Flasche, Rosalind M. Eggo

## Abstract

As the number of novel coronavirus cases grows both inside and outside of China, public health authorities require evidence on the effectiveness of control measures such as thermal screening of arrivals at airports. We evaluated the effectiveness of exit and entry screening for 2019-nCoV infection. In our baseline scenario, we estimated that 46.5% (95%CI: 35.9 to 57.7) of infected travellers would not be detected, depending on the incubation period, sensitivity of exit and entry screening, and the proportion of cases which are asymptomatic. Airport screening is unlikely to detect a sufficient proportion of 2019-nCoV infected travellers to avoid entry of infected travellers. We developed an online tool so that results can be updated as new information becomes available.

## Background

As of 29 Jan 2019, 5,997 confirmed and 9,239 suspected cases of 2019-nCoV have been reported from China with 132 deaths confirmed so far ^1^. There were cases in at least 15 other countries, identified due to symptoms plus recent travel history to Hubei province, China, which strongly suggests that the reported cases constitute only a small fraction of the actual number of infected individuals in China ^2^. While the most affected region, Hubei province, has now ceased air travel and closed major public transport routes ^3^ it is unlikely that these measures will fully contain the outbreak.

Despite limited evidence for its effectiveness ^4,5^, airport screening is frequently implemented to limit the probability of infected cases entering other regions or countries. Here we use the available evidence to evaluate the effectiveness of exit and entry screening for detecting travellers with 2019-nCoV. We also provide an online app so that results can be updated as new information becomes available.

### Simulation of travellers at each stage of infection

We simulated 100 infected air travellers who are still infected with 2019-nCoV on arrival at their destination and hence would pose a risk for seeding transmission in a new region. The duration of travel is the flight time plus a small amount of additional travel time for airport procedures. We assumed that most individuals will develop symptoms, including fever, at the end of their incubation period and progress to more severe symptoms after a few days, resulting in hospitalisation and isolation. Individuals may also exhibit an asymptomatic (subclinical) infection where they do not exhibit symptoms that would be detected by thermal scanning or cause them to seek medical care, although these individuals may be infectious. Travellers may exhibit severe symptoms during their travel; upon arrival they are hospitalised without undergoing entry screening. We then estimated the proportion of travellers who would be detected by exit and entry screening, develop severe symptoms during travel, or go undetected, under varying assumptions of:

- the proportion of asymptomatic infections;
- the sensitivity of exit and entry screening;
- the duration of travel;
- the incubation period;
- the time from the start of symptoms until hospitalisation (Table 1).

**Table 1:** Parameter values and assumptions for the baseline scenario

| Parameter | Value (baseline scenario) | Source |
| --- | --- | --- |
| Travel time | 12 hours | Beijing - London <sup>9</sup> |
| Sensitivity of exit screening | 86% | 86% for Infrared thermal image scanners <sup>10</sup> |
| Sensitivity of entry screening | 86% | 86% for Infrared thermal image scanners <sup>10</sup> |
| Proportion of asymptomatic infections undetectable by typical screening procedures | 17% | 1 of 6 reported asymptomatic in a 2019-nCoV family cluster <sup>11</sup> |
| Incubation period | Gamma distributed, mean 5.2 days, variance 4.1 days | Reported mean, variance estimated from uncertainty interval of mean<sup>1</sup> |
| Time from symptom onset to hospitalisation | Gamma distributed, mean 9.1 days, variance 14.7 days | Reported mean, variance estimated from uncertainty interval of mean<sup>1</sup> |

We assume that the time of starting travel is randomly and uniformly distributed between the time of infection and twice the expected time to severe disease; however, we only consider those travellers who depart before their symptoms worsen to the point that they would seek hospital care ^6^. We simulate travellers with individual incubation period, time from onset to severe disease, flight start times and detection success at exit and entry screening according to the screening sensitivities (Figure 3). We consider only the travel of infected individuals (not the proportion of travellers who are infected) and therefore assume a screening specificity of 100%. An individual will be detected at exit screening if their infection is not asymptomatic, their departure time exceeds their incubation period, and their stochastic exit screening success indicates detection. An individual will be detected at entry screening if their infection is not asymptomatic, their incubation period ends after their departure but before their arrival, they have not been detected at exit screening, and their entry screening success indicates detection. Entry screening detections are further divided into detection due to severe symptoms and detection of mild symptoms via equipment such as thermal scanners. We used 10,000 bootstrap samples to calculate 95% confidence intervals.

**Figure 1:**
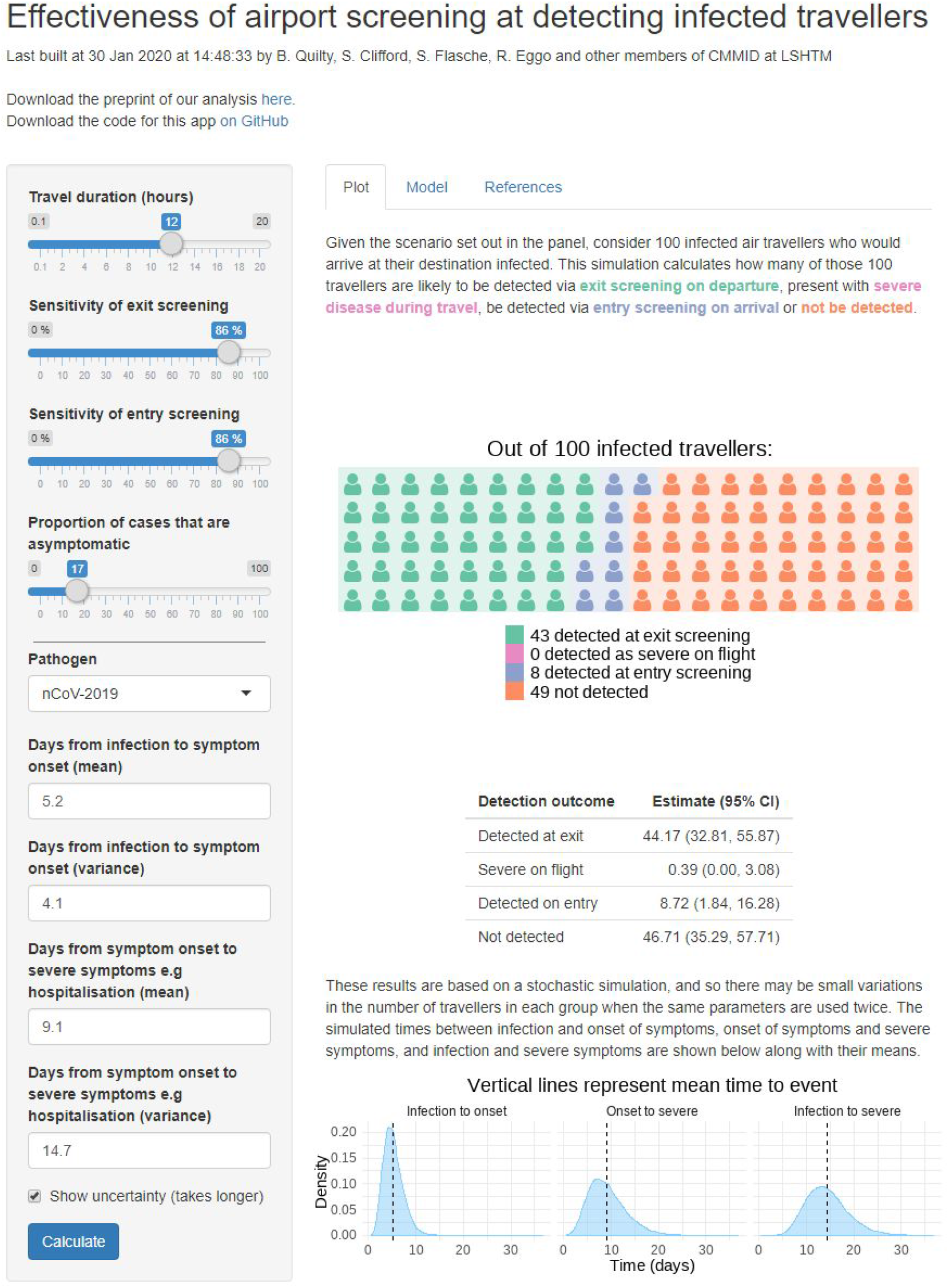
Screenshot of app ^7^ displaying the number of infected travellers detected for the baseline assumptions (Table 1) with 95% bootstrap confidence intervals and the time distributions for incubation period and time to severe disease e.g hospitalisation. Results are from stochastic simulation, and so there may be small variations in the number of travellers in each group when the same parameters are used twice. Sliders are provided to modify the duration of travel, the sensitivity of both exit and entry screening, the proportion symptomatic, and the natural history parameters for the infection.

**Figure 2:**
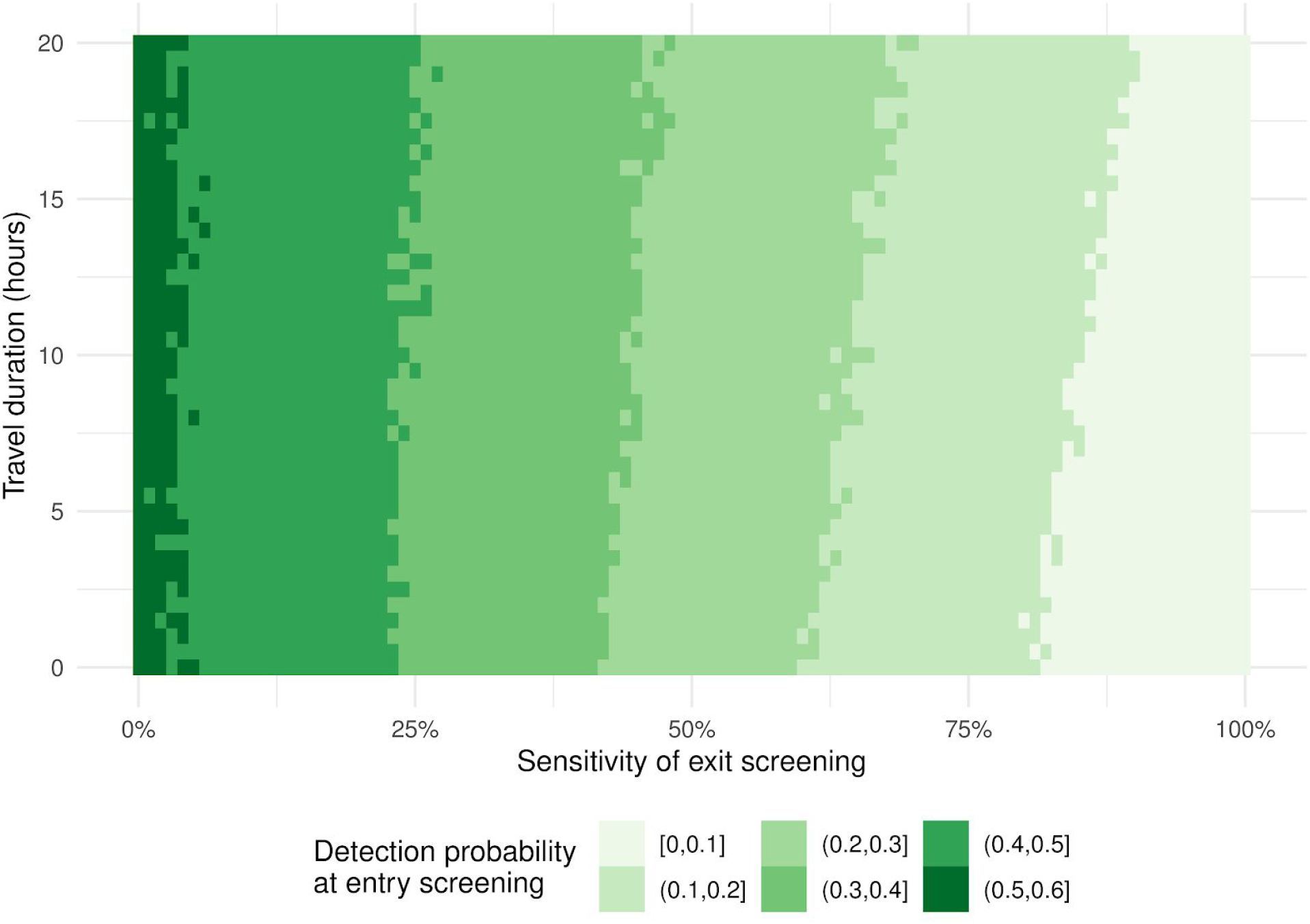
Probability of detecting infected travellers at entry screening for varying travel durations and sensitivities of exit screening. Each cell is a mean of 10,000 model simulations. Other parameters (Incubation period, symptom onset to hospitalisation period, and asymptomatic rate) were fixed at baseline assumptions (Table 1). Intervals are probabilities of detection, binned at increments of 10% (0-10%, 10-20%, etc.).

**Figure 3:**
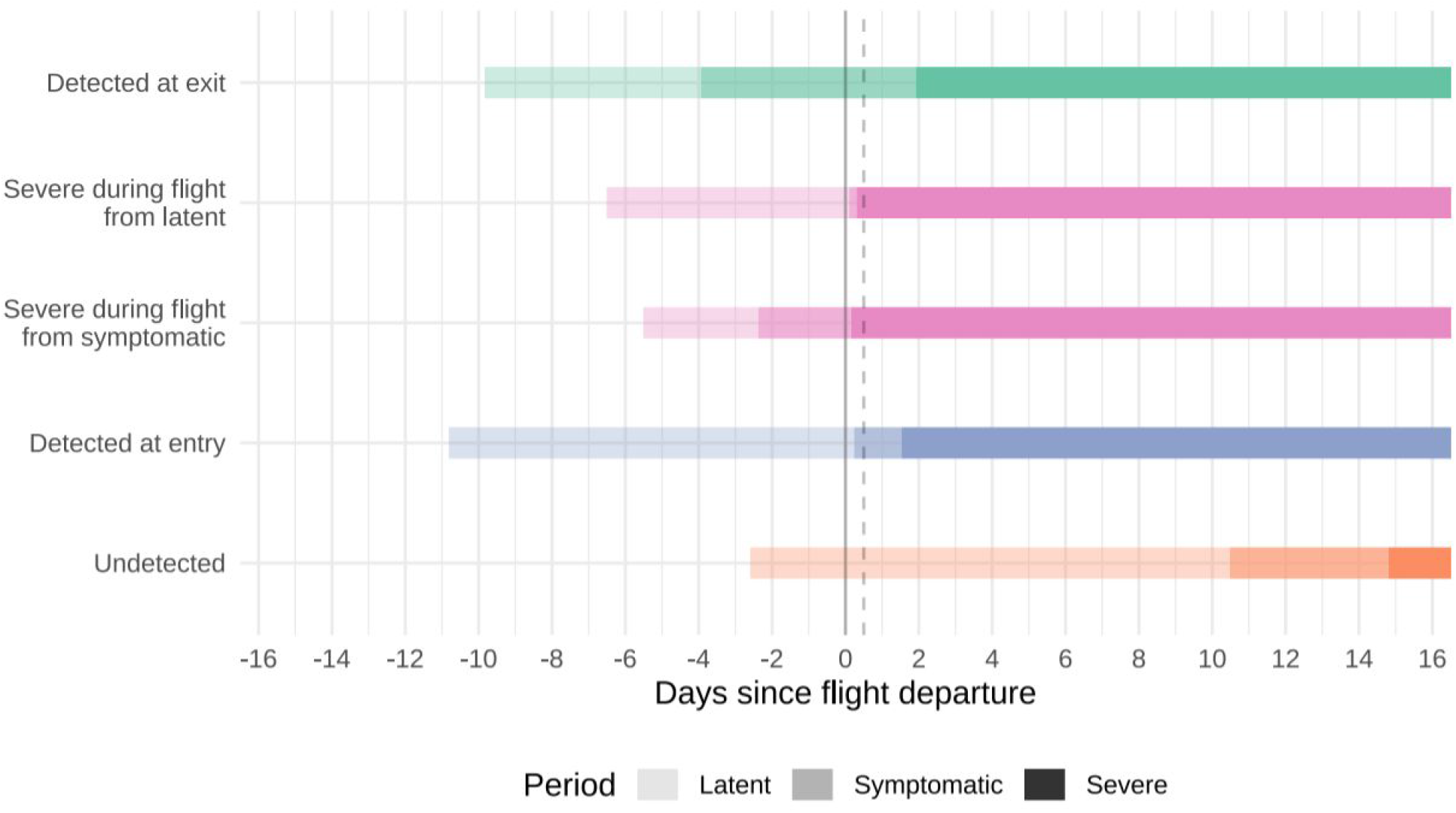
Simulated infection histories of infected travellers. The latent period begins on infection and travellers then progress to symptomatic and severe symptoms. Travellers may fly at any point within the latent or symptomatic phases; any would-be travellers who show severe symptoms are hospitalised prior to exit. Vertical lines represent the exit screening at start of travel (solid) and entry screening at end of travel (dashed) 12 hours later.

The model code is available via GitHub ^7^ and the results can be further explored in an R Shiny app at https://cmmid-lshtm.shinyapps.io/traveller_screening/ (Figure 1).

### Effect of screening on detection

For the baseline scenario we estimated that 44 (95%CI: 33-56) of 100 infected travellers would be detected by exit screening, 0 (95% CI: 0-3) case would develop severe symptoms during travel, 9 (95%CI: 2-16) additional cases would be detected by entry screening, and the remaining 46 (95%CI: 36-58) would not be detected.

The effectiveness of entry screening is largely dependent on the effectiveness of exit screening in place. Under baseline assumptions, entry screening could detect 53 (95%CI: 35-72) instead of 9 infected travellers if no exit screening was in place. However, the probability of developing symptoms mid-flight increases with flight time and hence exit screening is more effective for longer flights (Figure 2).

Syndromic screening designed to prevent infected and potentially infectious cases entering undetected is highly vulnerable to asymptomatic infections and long incubation periods. If the baseline scenario is modified to have 0% asymptomatic infections and 100% sensitivity of entry screening, the incubation period will need to be approximately 10-fold shorter than the period from symptom onset to severe disease (e.g. hospitalisation) in order to detect more than 90% of infected travellers that would not otherwise report illness at either exit or entry screening.

## Conclusions

*A*s a response to the ongoing outbreak of a novel coronavirus originating in Wuhan, the Chinese government has implemented exit screening for international flights leaving China ‘s major airports. Thermal scanning, which can identify passengers with fever (high external body temperature), allows for passengers exhibiting symptoms of 2019-nCoV infection to be tested before they board a flight. Similarly, entry screening for flights originating in the most affected regions may be considered at airports in regions in and outside China. We estimate that the key goal of syndromic screening at airports - to prevent infected travellers from entering regions with little or no ongoing transmission - is only achievable if the rate of asymptomatic infections that are transmissible is negligible, screening sensitivity is almost perfect, and the incubation period is short. Based on data from Li et al. (2020)^8^, 2019-nCoV has a shorter incubation period than SARS, and a higher rate of asymptomatic infections^9^. Under generally conservative assumptions on sensitivity, we find that 46.5% of infected travellers will enter undetected.

Entry screening is an intuitive barrier for the prevention of infections entering a region or country. However, evidence on its effectiveness is limited and given the lack of specificity, it generates a high overhead of screened travellers uninfected with the targeted pathogen^4^. For example, when entry screening was implemented in Australia in response to the 2003 SARS outbreak, 1.84 million people were screened, 794 were detained, and no cases were confirmed^10^. While some cases of 2019-nCoV have been identified through airport screening in the current outbreak we estimate that likely many more infected travellers have passed through screening undetected.

It is important to note that our estimates are based on a number of key assumptions that cannot yet be informed directly by evidence from the ongoing 2019-nCoV outbreak. The current outbreak has spread rapidly and early evidence suggests that the average severity is lower than SARS. This may also suggest a substantial proportion of asymptomatic cases. A recent analysis of a family transmission cluster is based on a small sample size but 1 in 6 infections was asymptomatic ^9^; a major impediment for the effectiveness of syndromic screening. However, if asymptomatic cases were not infectious they would not pose a risk for seeding infection chains on arrival. To allow easy adaptation of our results as new insight becomes available in the coming weeks we developed an online app.

Furthermore, the most up-to-date data on the incubation period or the time until recovery of 2019-nCoV has been used in this analysis, yet these figures are likely to change over time as more data becomes available. Unless the incubation period is only a small fraction of the duration of clinical symptoms, and fever in particular, syndromic screening is likely to detect an insufficient fraction of infected cases to prevent local infections. In addition, the sensitivity of airport screening for the detection of 2019-nCoV has not been evaluated. However, we chose conservative estimates and show that with reduced sensitivity the effectiveness of syndromic screening would further decline.

In many international airports, information is provided to travellers from affected regions advising recommended action if they develop symptoms on or after arrival^11–14^. While this is likely to be an effective strategy, we find that airport screening for initial symptoms, via thermal scanners or similar, on either exit or entry is unlikely to detect a sufficient proportion of 2019-nCoV infected travellers in order to avoid entry of infected travellers and therefore the potential for seeding of local transmission.

## Data Availability

All data is available on the referenced webpage.

https://cmmid-lshtm.shinyapps.io/traveller_screening/

https://github.com/bquilty25/airport_screening

